# Neutralizing antibody against SARS-CoV-2 spike in COVID-19 patients, health care workers and convalescent plasma donors: a cohort study using a rapid and sensitive high-throughput neutralization assay

**DOI:** 10.1101/2020.08.02.20166819

**Authors:** Cong Zeng, John P. Evans, Rebecca Pearson, Panke Qu, Yi-Min Zheng, Richard T. Robinson, Luanne Hall-Stoodley, Jacob Yount, Sonal Pannu, Rama K. Mallampalli, Linda Saif, Eugene Oltz, Gerard Lozanski, Shan-Lu Liu

**Author notes:** To whom correspondence should be addressed: Dr. Shan-Lu Liu, The Ohio State University, Columbus, OH 43210, USA. Equal contribution (Cong Zeng, John P. Evans).

## Abstract

Rapid and specific antibody testing is crucial for improved understanding, control, and treatment of COVID-19 pathogenesis. Herein, we describe and apply a rapid, sensitive, and accurate virus neutralization assay for SARS-CoV-2 antibodies. The new assay is based on an HIV-1 lentiviral vector that contains a secreted intron Gaussia luciferase or secreted Nano-luciferase reporter cassette, pseudotyped with the SARS-CoV-2 spike (S) glycoprotein, and is validated with a plaque reduction assay using an authentic, infectious SARS-CoV-2 strain. The new assay was used to evaluate SARS-CoV-2 antibodies in serum from individuals with a broad range of COVID-19 symptoms, including intensive care unit (ICU) patients, health care workers (HCWs), and convalescent plasma donors. The highest neutralizing antibody titers were observed among ICU patients, followed by general hospitalized patients, HCWs and convalescent plasma donors. Our study highlights a wide phenotypic variation in human antibody responses against SARS-CoV-2, and demonstrates the efficacy of a novel lentivirus pseudotype assay for high-throughput serological surveys of neutralizing antibody titers in large cohorts.

## INTRODUCTION

The COVID-19 pandemic, caused by the SARS-CoV-2 virus, which currently engulfs the globe, has led to over 10 million cases and 500,000 deaths world-wide *(1, 2)*. Without an effective vaccine, significant public health measures are required to minimize the spread of the virus, including masks and social distancing complemented with wide-spread testing and contact tracing. Reliable and easily deployed testing is necessary to fully understand the spread of the virus. Among these, antibody assays are a critical arm of our testing capacity that will allow for large-scale serum surveillance and closer monitoring of immunity in individual patients *(3-6)*. Even after the development of a vaccine, large-scale antibody testing is needed to track vaccine and herd immunity as SARS-CoV-2 infections subside *(7, 8)*. This is particularly important since antibody responses to SARS-CoV-2 infection may wane over time, as is the case for other human coronaviruses *(9-11)*. Additionally, in the short-term, convalescent plasma from recovered SARS-CoV-2 patients has been used to treat severe COVID-19 cases, and testing convalescent plasma for high SARS-CoV-2 neutralizing antibody titers can improve this treatment strategy *(12-15)*. Thus, accurate determinations of neutralizing antibody titers are of particular importance for both surveillance and testing of patient serum.

Recent evidence has suggested that several available antibody tests exhibit poor correlation with neutralizing titers *(4)*. Following the large SARS-CoV-2 outbreak in New York, several studies investigated the seroconversion of previously infected individuals *(4-6)*. The vast majority of COVID-19 patients were shown to seroconvert by enzyme-linked immunosorbent assays (ELISAs) *(5)*. However, other analyses have demonstrated that recovered SARS-CoV-2 patients often have only weak titers of neutralizing antibodies against the virus *(4, 6, 16, 17)*, further emphasizing the need for reliable antibody neutralization tests.

Several pseudotype virus neutralizing antibody tests have been developed for SARS-CoV-2 *(18-22)*. To avoid the need for live virus and a BSL3 facility, these assays typical utilize an HIV- or VSV-based vector pseudotyped with the SARS-CoV-2 S protein *(4, 20, 23)*. This approach allows for the accurate detection of anti-S neutralizing antibodies in a more commonly available BLS2 facility. However, existing neutralization assays utilize a firefly luciferase, Renilla luciferase, or green fluorescent protein (GFP) reporter cassette that typically require a cell-lysis step, increasing the experimental timeline and reducing the scale-up capacities. Here, we developed a SARS-CoV-2 antibody neutralization assay that utilizes an HIV-1 vector bearing a secreted intron Gaussia or Nano-Luciferase reporter cassette and is pseudotyped with SARS-CoV-2 S protein. This approach permits sensitive, rapid, and accurate testing of neutralizing antibody titers without a cell-lysis step, and was validated with an authentic SARS-CoV-2 USA-WA-1 strain in plaque reduction virus neutralization assays (PRVN). Using our S pseudotype virus assay, we examined neutralizing antibody titers for 221 blinded serum samples, which include 104 hospitalized COVID-19 patients (49 ICU patients and 55 inpatients), 42 OSU health care workers, 38 convalescent plasma donors - all are RT-PCR confirmed, and 37 negative control samples collected prior to September, 2019. Our neutralization results are highly concordant with that of SARS-CoV-2 nucleocapsid (N)-based IgG antibody ELISA measurements. Prior reports are conflicting on the presence of cross-reactive antibodies to SARS-CoV and SARS-CoV-2 *(24-27)*. In the patient samples tested, we found no antibody cross-neutralization of SARS-CoV in COVID-19 patients. Additionally, our results bolster prior reports that more severe COVID-19 cases tend to have higher neutralizing antibody titers *(25)*, but also indicate that health care workers, as well as convalescent plasma donors, have a varied neutralizing antibody response, which will require more attention from the scientific and public health communities.

## RESULTS

### Generation of secreted intron Gaussia luciferase-based lentiviral pseudotypes bearing SARS-CoV-2 spike

The S protein of coronavirus can pseudotype a variety of viral vector systems, including lentiviral vectors *(18-20)*. Differing from most lentiviral pseudotypes, which bear GFP, alkaline phosphatase, or firefly/renilla/nanoLuc luciferases as a reporter, we chose gaussian luciferase (Gluc) as the transducing gene, given its natural secretion into mammalian cell culture media and high sensitivity, thus facilitating detection *(28)*. To prevent Gluc activity in producer cells, which may cause a high background signal in the media of target cells, we took advantage of a published intron Gluc (inGluc) system *(29, 30)*. In this lentiviral vector, the anti-sense Gluc reporter gene is interrupted by an intron oriented in the sense direction of the HIV-1 NL4.3 genome. As such, expression of Gluc can be detected only in vector-transduced target cells after infection (**Fig. 1A**).

**Figure 1.**
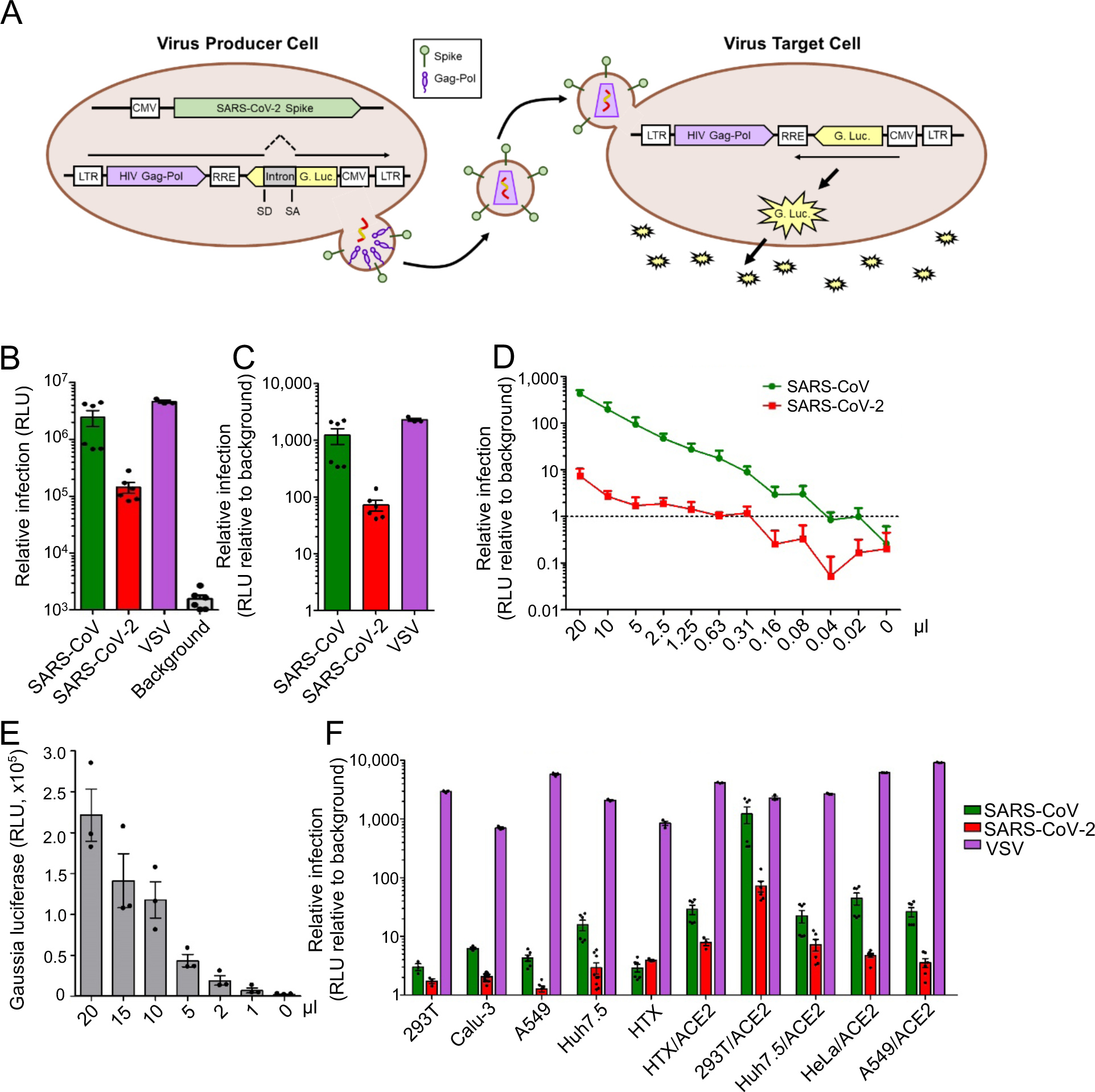
Intron-Gluc-based HIV-1 lentiviral S pseudotypes bearing SARS-CoV-2 spikes. 293T cells seeded on 6-well plates were co-transfected with 0.8 μg HIV-1-NL4.3-inGluc vector plus 0.4 μg SARS-CoV-2 spike-coding plasmid; 48 h post-transfection, viral supernatant was harvested and used to infect target cells. Unless otherwise indicated, 293T/ACE2 cells were used for infection. **(A)** Schematic representation of the pseudoviral production and infection. Note that Gluc activity can only be detected in virus-infected target cells, but not in the virus-producing cells because of the presence of an intron inserted in the sense of the vector that splits the Gluc gene into two parts. **(B and C)** Titers of HV-1 inGluc pseudotypes bearing the spikes of SARS-CoV, SARS-CoV-2 or VSV-G; absolute luciferase readouts (B, n=6), and relative infectivity compared to the background (C, n=6) were plotted, respectively. **(D)** Indicated doses of viral supernatant were used to infect 293T/ACE2 cells seeded in 24-well plates, and 20 μL of supernatant of virus-infected cells were used to measure the Gluc activity as shown. The dashed line indicates the background of luciferase activity. **(E)** Indicated amounts of culture media harvested from virus-infected cells were used to measure Gluc activity (n=3). **(F)** Relative infectivity of HIV-inGluc pseudotypes bearing S proteins of SARS-CoV, SARS-CoV-2 or VSV-G in indicated target cells, with parental or those overexpressing ACE2.

To test the HIV-1 NL4.3-inGluc vector for SARS-CoV-2, we transfected 293T cells seeded on a 6-well plate, which also expresses HIV-1 Gag-Pol but not Env, together with a plasmid encoding the S protein of SARS-CoV-2 that is tagged with C9 (TETSQVAPA) at the C-terminus to facilitate detection *(31)*. In parallel, the S of SARS-CoV, which also has a C9 tag, or VSV-G, were co-transfected. Supernatants from transfected cells were harvested, and used to infect target 293T cells overexpressing the SARS-CoV-2 receptor ACE2 (292T/ACE2) *(32, 33)* in 96-well plates, and Gluc activity was measured at 24, 48 and 72 h post-infection (**Fig. 1B**). We found that SARS-CoV-2 S protein can pseudotype the inGluc-based lentiviral vector, although the infectivity of SARS-CoV-2 S pseudotypes was 5∼10-fold lower than that of SARS-CoV; VSV pseudotypes exhibited the highest titer as would be expected (**Fig. 1B**). The relative infectivity of SARS-CoV, SARS-CoV-2 and VSV pseudotypes in 293T/ACE2 cells, as compared to the background control, was plotted in **Fig. 1C**, showing that lentiviral SARS-CoV-2 S pseudotypes consistently produced a luciferase signal 50∼100-fold above background at 2-3 days post-infection.

To gain insight into the efficiency of SARS-CoV-2 S pseudotyping and the sensitivity of this lentiviral system, we applied different amounts of viral stocks in 2-fold serial dilutions, i.e., 20, 10, 5, 2.5…0.02 μl, to target cells (**Fig. 1D**), along with serially reduced volumes of culture media of infected cells, i.e., 20, 15, 10, 5, 2, 1 μl, to measure Gluc activity (**Fig. 1E)**. Activity can be detected even with less than 1 μl of culture media for Gluc measurements, and by calculating the minimal amount of virus stock and culture media that gave a Gluc signal above background, we were able to estimate that the infectious units (IU) of SARS-CoV-2 and SARS-CoV lentiviral Gluc pseudotypes were ∼8.0 × 10^4^ and ∼3.2 × 10^5^ per ml,respectively.

To identify the best target cell line for our lentiviral system, we compared the infectivity of HIV-NL4.3-inGluc bearing SARS-CoV-2 S, along with that of SARS-CoV S and VSV-G, in a panel of cell lines, some of which were engineered to overexpress ACE2. Among these cells, we found that 293T/ACE2 cells consistently offered the most robust and consistent Gluc signals (**Fig. 1F**), likely because of their highly proliferative nature compared to other cell lines. Accordingly, 293T/ACE2 cells were used in the experiments below.

### Processed SARS-CoV-2 spike protein is incorporated into the lentiviral vector

The S constructs used above for pseudotyping the HIV-NL4.3-inGluc vector contain C9 tags at their C-termini, which allowed us to determine and compare their expression in cells and in viral particles. Moreover, it is important to know the pseudotyping efficiency of C9-tagged spikes versus their corresponding wildtype (WT) counterparts. As shown in Fig. 2A, the titer of the C9-S-bearing SARS-CoV-2 had a 5∼10-fold-increased titer relative to the WT counterpart; however, C9-spike-bearing SARS-CoV did not exhibit a difference from its WT, at least in the 293T/ACE2 cells. We then performed western blotting to examine the expression of the S proteins in viral producer cells. We found that the C9-tagged SARS-CoV and SARS CoV-2 S proteins were expressed, with somewhat similar efficiency, although the S protein of SARS-CoV-2 was cleaved more efficiently compared to that of SARS-CoV (**Fig. 2B**). Western blotting analysis of purified S-pseudotyped viral particles revealed that the C9-S protein of SARS-CoV was more efficiently incorporated into HIV-1 lentiviral particles compared to that of SARS-CoV-2. Despite this, the latter was present in its processed form (**Fig. 2C**).

**Figure 2.**
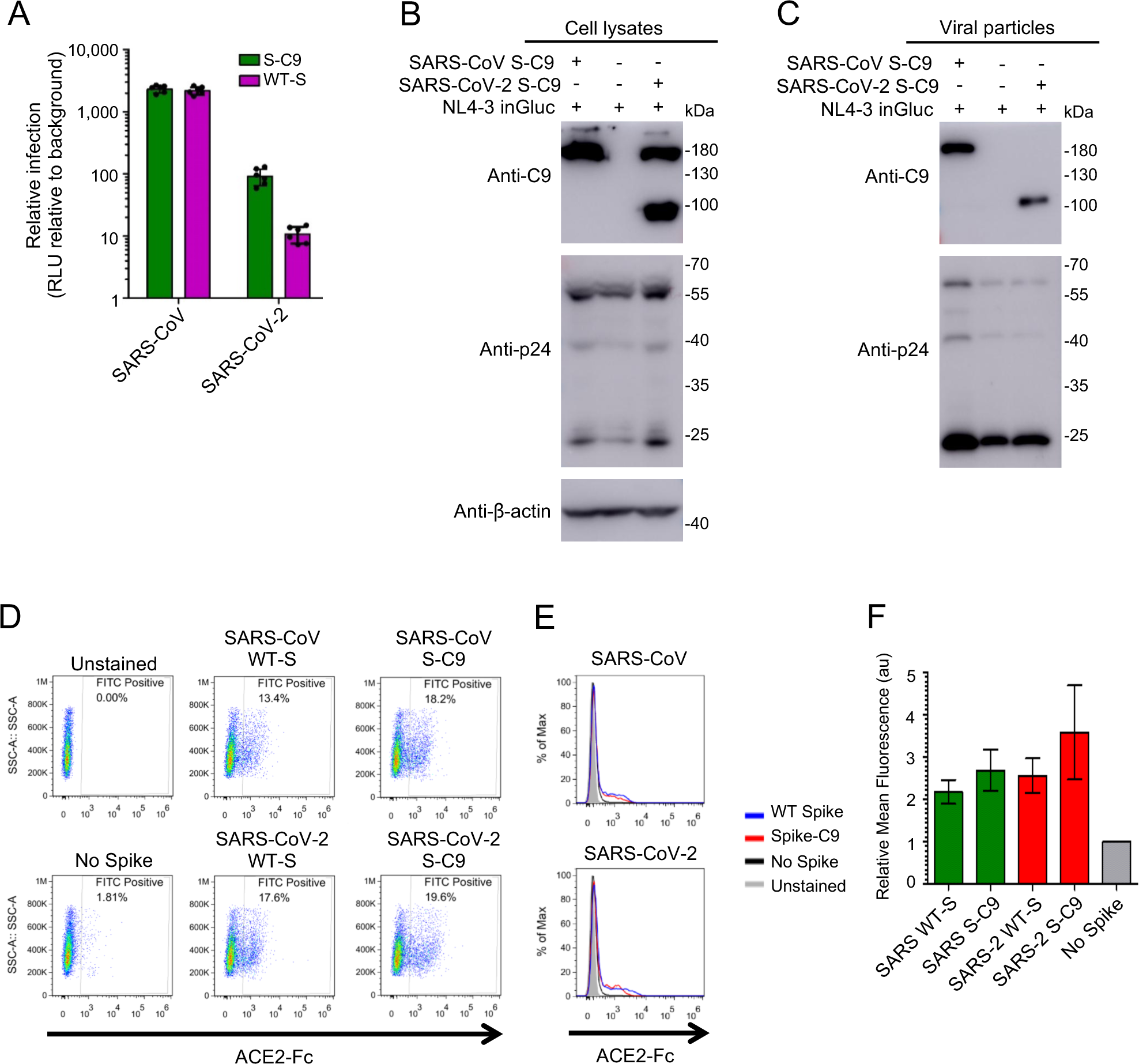
Comparison of HIV-1 inGluc pseudotypes bearing C9-tagged spikes of SARS-CoV or WTs. **(A)** Relative infectivity. Experiments were performed as described as Figure 1 B and C, except that either WT or C9-tagged spikes were used for virus production. **(B and C)** Western blotting analysis of C9-tagged S protein expression in the virus-producing cells (B) and purified viral particles (C). Viral production was carried as described in Figure 1, and viral producer cells were lysed and analyzed by Western blotting using anti-C9, anti-p24 and/or anti-β-actin, respectively. **(D, E and F)** Virus-producing cells were digested with PBS-5 mM EDTA, and incubated with 10 ug/ml sACE2-Fc for 2 h; cells were washed 3 times, incubated with FITC-labeled anti-human Fc for 45 min, and were analyzed by flow cytometry. **(D)** Representative cell populations analyzed for SARS-CoV and SARS-CoV-2; the percent of positive cells for FITC-anti-human Fc was shown. **(E)** Histogram analysis of virus-producing cells for SARS-CoV and SARS-CoV-2. **(F)** Relative mean fluorescence intensities of cells expressing indicated spikes. Note that cells transfected with an empty vector pCIneo served as negative control, the fluorescence intensity of which was set to 1.

Due to the lack of a common antibody allowing us to detect all forms of S proteins (WT and C9 for both SARS-CoV and SARS-CoV-2), we used a soluble form of ACE2-human Fc fusion protein (ACE2-hFc) and anti-human Fc in flow cytometry to examine the expression of these S proteins on the surface of the viral producer cells. As shown in **Fig. 2D, E and F**, no significant differences were observed between these spikes, regardless of whether they were SARS-CoV, SARS-CoV-2 or C9-tagged variants. Altogether, these results demonstrate that the S glycoprotein of both SARS-CoV-2 and SARS-CoV can pseudotype the lentiviral vector, with SARS-CoV-2 exhibiting a lower pseudotyping efficiency than SARS-CoV. However, the presence of a C9 tag on the C-terminus of SARS-CoV-2 S substantially increases the titer of lentiviral pseudotypes bearing SARS-CoV-2 S.

### Application of inGluc-based lentiviral SARS-CoV-2 S pseudotypes for detecting SARS-CoV-2/COVID-19 neutralizing antibody and correlations with infectious virus-based plaque reduction

One goal of developing sensitive and convenient lentiviral pseudotypes bearing SARS-CoV-2 S is to evaluate the neutralizing antibody response in COVID-19-confimed cases and SARS-CoV-2-exposed individuals. Towards this goal, we initially tested a small group of eight blinded patient serum samples, with limited dilutions. Briefly, the samples were pretreated by heat-inactivation at 56°C for 1 h, serially diluted at 1:20, 40, 80 and160, and then incubated with inGluc SARS-CoV-2 S pseudotypes at 37°C for 1 h. The virus-sera mixtures were subsequently added to 293T/ACE2 cells to allow infection for 6 h before being removed from target cells; culture media were collected from cells at 24 and 48 h post-infection and measured for Gluc luciferase activity. As shown in **Fig. 3A**, samples 3 and 8 exhibited strong neutralizing activity, having > 90% inhibition of viral infectivity at 1:160 compared to samples 2 and 5, which showed 50% inhibition of viral infectivity between 1:80 and 1:160. In sharp contrast, negative control samples 4, 6 and 7, which were collected prior to September, 2019, showed only background level of inhibition. Importantly, the neutralization pattern of patient sample 8 using WT S-bearing inGluc pseudotypes was identical to that using the S-C9-bearing pseudotypes (**Fig. 3A**), indicating that the C9-tagged SARS-CoV-2 S faithfully mimics that of WT S in the neutralization assay. The virus neutralization results were consistent with nucleocapsid (N)-based antibody ELISA OD_450_ values, with samples 2, 3, 5 and 8 having a range between 0.616 and 1.216 in contrast to samples 1, 2, 6 and 7, which were 0.125 to 0.238 (the ELISA cut-off was set to ∼0.40) (**Fig. 3B**). Results from this initial blinded testing gave us confidence that the inGluc-based SARS-CoV-2 S-pseudotype virus neutralization assay is potentially useful for determining the neutralizing antibody titer in COVID-19 patients.

**Figure 3.**
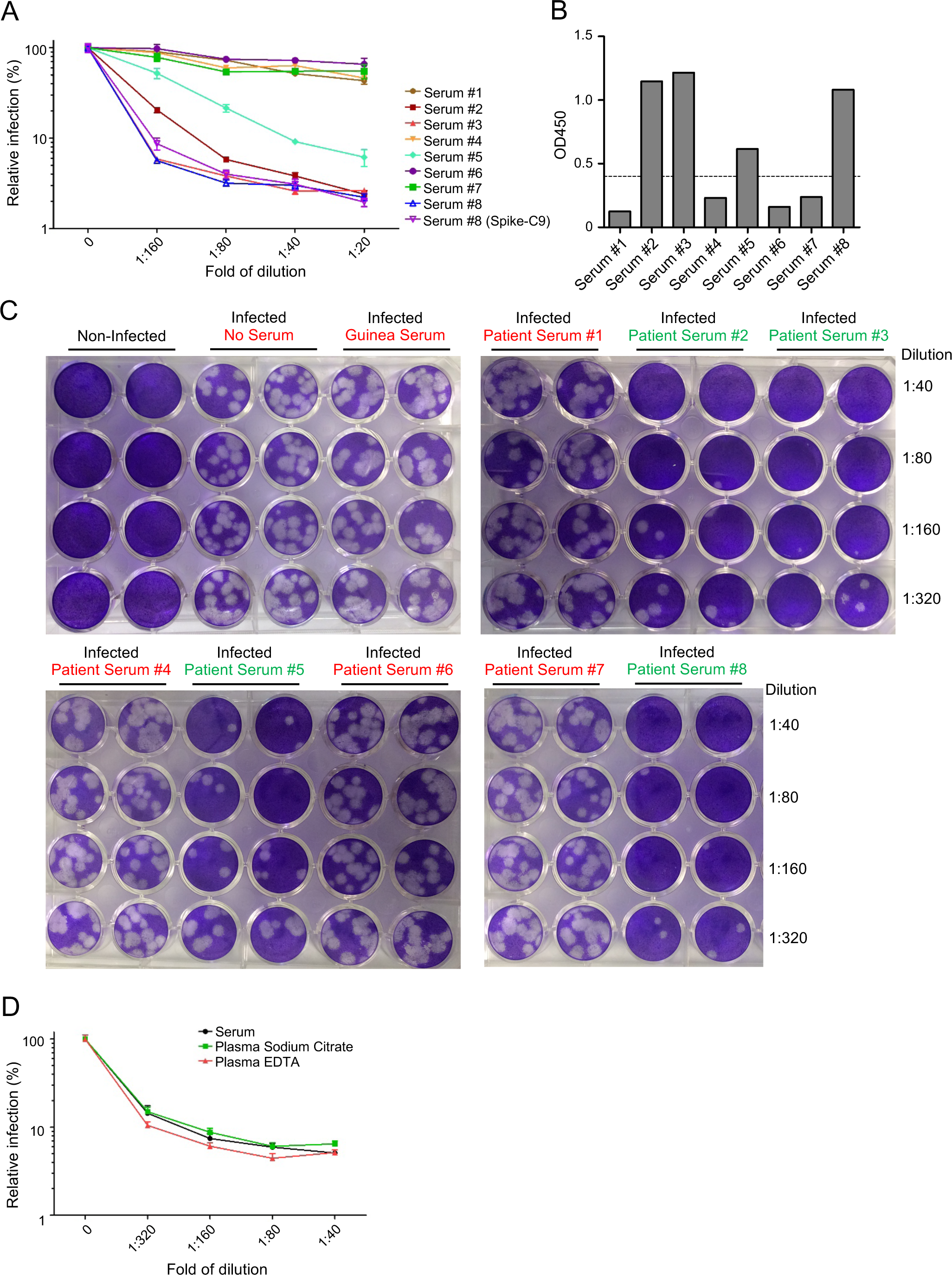
Neutralization of SARS-CoV-2 by COVID-19 patient sera—validation of inGluc-based lentiviral pseudotypes using an authentic SARS-CoV-2 US-WA-1 strain. Note that all samples tested here and throughout the studies were blinded before testing. **(A)** A group of 8 blinded patient sera was tested for neutralization of the SARS-CoV-2 lentiviral S pseudotypes bearing C9-tagged SARS-CoV-2 Sor WT. Note that only patient serum sample (#8) was tested for the WT spike, the pattern of which almost perfectly overlaps with that of C9-tagged spike. **(B)** The ELISA OD_450_ values of 8 samples are presented (cut-off: 0.40 – dashed line). **(C)** Results of infectious SARS-CoV-2 plaque-reduction neutralization assay for testing of 8 blinded samples. Vero-E6 cells were infected for 3 days with infectious SARS-CoV-2, pre-treated with or without the indicated diluted sera. Cells were fixed and stained with 0.25% crystal violet for visualization of plaques. Note that the BEI guinea pig antiserum to SARS-CoV did not inhibit SARS-CoV-2 infection. **(D)** Types of serum or plasma samples did not appear to affect the neutralization pattern generated by inGluc-based lentiviral pseudotypes bearing SARS-CoV-2 spike. Serum, sodium citrate-treated plasma, and EDTA-treated plasma from the same patient were used for SARS-CoV-2 S pseudotype neutralization.

We further validated the inGluc-based virus neutralization assay with a plaque reduction virus neutralization (PRVN) assay using an infectious SARS-CoV-2 strain, USA-WA-1. As shown in **Fig. 3C**, the numbers of SARS-CoV-2 plaques were greatly reduced by serum samples 2, 3, 5 and 8, with samples 3 and 8 being stronger than samples 2 and 5 in reducing plaques. In contrast, serum samples 1, 4, 6 and 7 had no impact on plaque numbers, which were similar to no serum treatment. These results correlated completely with that of the S pseudovirus-based virus neutralization assay shown in **Fig. 3A**, although we were unable to calculate their exact titers due to limited sample volumes and dilutions. Of note, guinea pig serum against SARS-CoV (BEI Resources, Cat# NR-10361, deposited by Dr. Linda Saif) did not inhibit the plaque-forming activity of SARS-CoV-2, indicating that it has no cross-reactivity with SARS-CoV-2. Because all samples tested above were sera, we next evaluated if plasma, collected in different forms, could also be used in our S-bearing lentiviral neutralization assay. As shown in **Fig. 3D**, serum, plasma-sodium citrate, and plasma-EDTA from one patient yielded an almost completely overlapping neutralization patterns, especially serum and plasma-sodium citrate. Thus, three forms of patient samples can be used for the inGluc-based SARS-CoV-2 neutralization assay.

### Cross-reactivity of antibodies between SARS-CoV-2 and SARS-CoV using inGluc-based lentiviral S pseudotypes

Given the inconsistency of reports regarding cross-reactivity of SARS-CoV antibodies with SARS-CoV-2 *(24-26)*, we extended our analyses using S-pseudotyped viruses. COVID-19 patient serum samples 2, 3, 5 and 8 and negative serum sample 1 shown in **Fig. 3A**, were further serially diluted from 1: 40 to 1:2,560 and tested for neutralizing antibody titers. As shown in **Fig. 4A**, polyclonal rabbit (BEI, Cat#NRC-777) and the guinea pig antisera against SARS-CoV showed potent inhibition of SARS-CoV infection, as would be expected; human monoclonal CR3022 (BEI, Cat #52392) also inhibited SARS-CoV infection, albeit with a much lower efficiency. None of the human COVID-19 patient serum samples, or the negative control serum sample 1 collected prior to September 2019, or the mouse 2B04 monoclonal antibody against SARS-CoV-2 *(34)* showed any neutralizing activity against SARS-CoV. For SARS-CoV-2, we observed that the mouse SARS-CoV-2 monoclonal antibody 2B04 potently inhibited SARS-CoV-2 infection, with IC_50_ of 3.007 ng/ml (**Fig. 4B**). Consistent with data shown in **Fig. 3A**, COVID-19 serum samples 2, 3, 5 and 8 showed varying degrees of SARS-CoV-2 neutralization, with samples 3 and 8 having an IC_50_ titer of 1:2,639 and 1:2,792, respectively (**Fig. 4B**). Overall, these results demonstrate that the inGluc-based virus neutralization assay can reliably determine neutralizing antibody responses in COVID-19 patients, and that sera or antibodies of human, mouse, rabbit, guinea pig, or non-human primates against SARS-CoV or SARS-COV-2 do not strongly cross-react in our inGluc-based virus neutralization assays.

**Figure 4.**
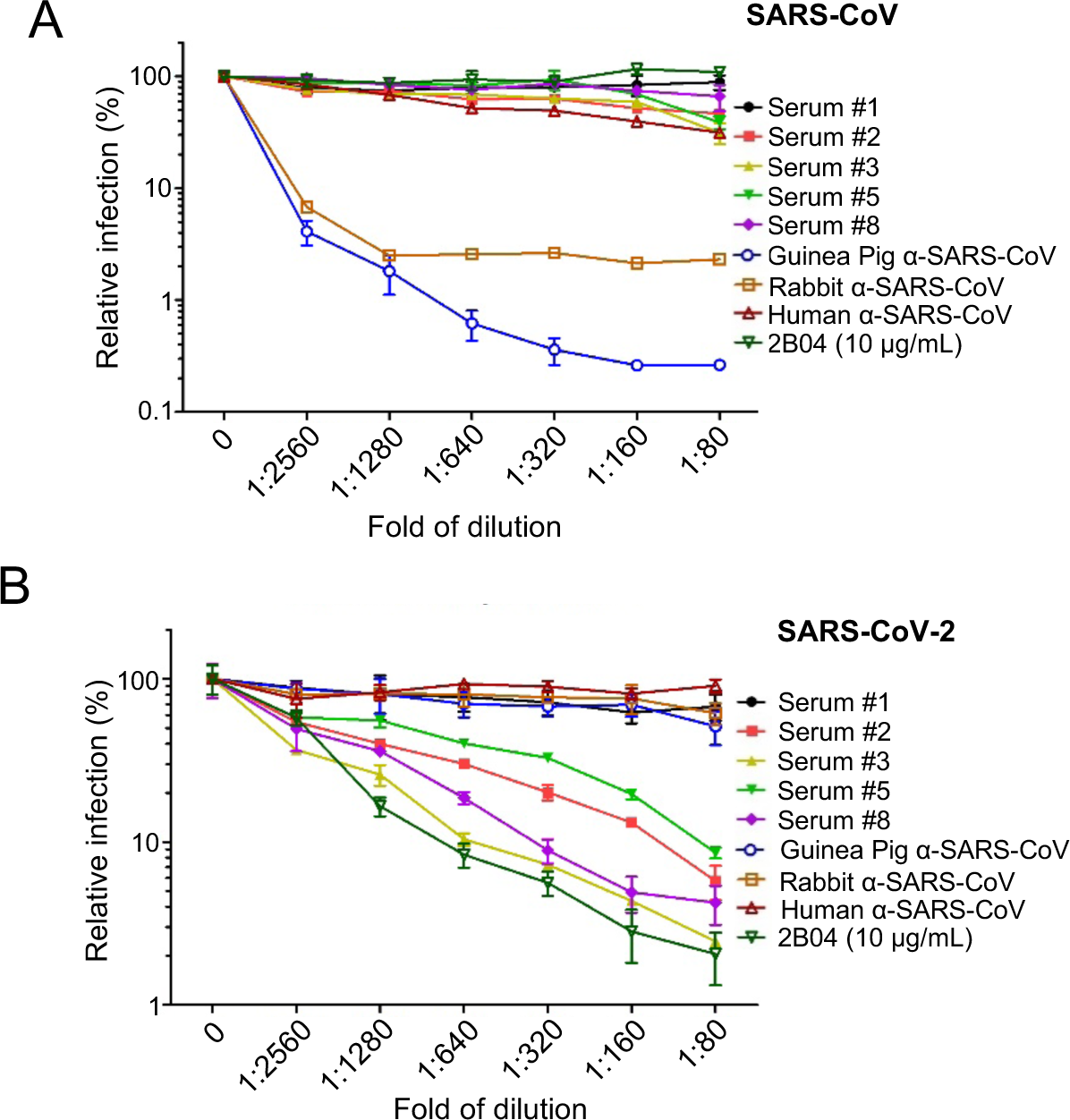
Examining the cross-reactivity of COVID-19 patient sera with SARS-CoV/SARS-CoV-2 S lentiviral pseudotypes. The neutralization assay was carried out as described in Figures 3 and 4, except that HIV-1 inGluc pseudotypes bearing SARS-CoV S **(A)** were used in parallel with that of SARS-CoV-2 **(B)**. In addition to 5 patient sera (samples 1, 2, 3, 5 and 8 from Fig. 3A), stocks of polyclonal (guinea pig and rabbit) or monoclonal (human) antibodies against SARS-CoV obtained from BEI were also tested; the exact concentration of these antibodies are unknown. A mouse monoclonal antibody 2B04 against SARS-CoV-2 (a kind gift from Ali Ellebedy), with a stock concentration of 10 μg/ml, was diluted to the same extent as the patient sera and other antibodies and tested.

### High-throughput testing of neutralizing antibody in hospitalized COVID-19 patients, health care workers and convalescent plasma donors

By expanding this assay to a 96-well high-throughput format, we determined the neutralizing antibody response in a cohort study. The cohort comprised 104 hospitalized patients (n=104), 55 of them were general hospitalized inpatients (n=55) and 49 were ICU patients (n=49); 42 Ohio State Medical Center health care workers (HCWs, n=42), all of which were PCR positive for SARS-CoV-2; as well as 38 blinded convalescent plasma donors (n=38). All of these samples were collected between April and May, 2020. In parallel, 37 frozen plasma samples, either from other respiratory disease cases or other patients collected prior to September, 2019, (n=37), were tested. We found that, in general, COVID-19 patients, including hospitalized and ICU patients, had high titers of SARS-CoV-2 neutralizing antibodies, with ∼20% (20 in 104) exhibiting a titer between 1:2,560 and 1:5,120, and with 12% (12 in 104) having a titer > 1:5,120 (**Figs. 5A, B and E**). Among these, ICU patients (∼45.0%; 22 in 49) showed an increased percentage of higher neutralizing titers above 1:1280 as compared to general hospitalized inpatients (∼36.0%, 20 in 55). We also noted that 14.5 % (8 in 55) of general hospitalized inpatients and 14.3 % of ICU patients (7 in 49) had no or low detectable neutralizing antibody detected (<1: 80), which was close to the ∼ 10% rate of previous reports (**Figs. 5A, B and E**). In sharp contrast, although all HCWs were confirmed RT-PCR+ for SARS-CoV-2 infection, ∼40% of the HCWs (17/42) were negative for SARS-CoV neutralizing antibody, and 36% (15/42) showed an intermediate titer between 1:80-320, with only 5% (2/42) having a titer over 1:1280 (**Fig. 5C and E)**. Notably, >55% of blinded convalescent plasma donor samples (21/38) exhibited a titer lower than 1:160 (**Fig. 5D and E)**, indicating that more than half of the blood donors did not qualify as convalescent donors for treatment of COVID-19 patients as per FDA guidelines *(35)*. Nevertheless, ∼26% of donor samples had a neutralization titer above 1:320, with one greater than 1:5,120 (**Fig. 5D and E**).

**Figure 5.**
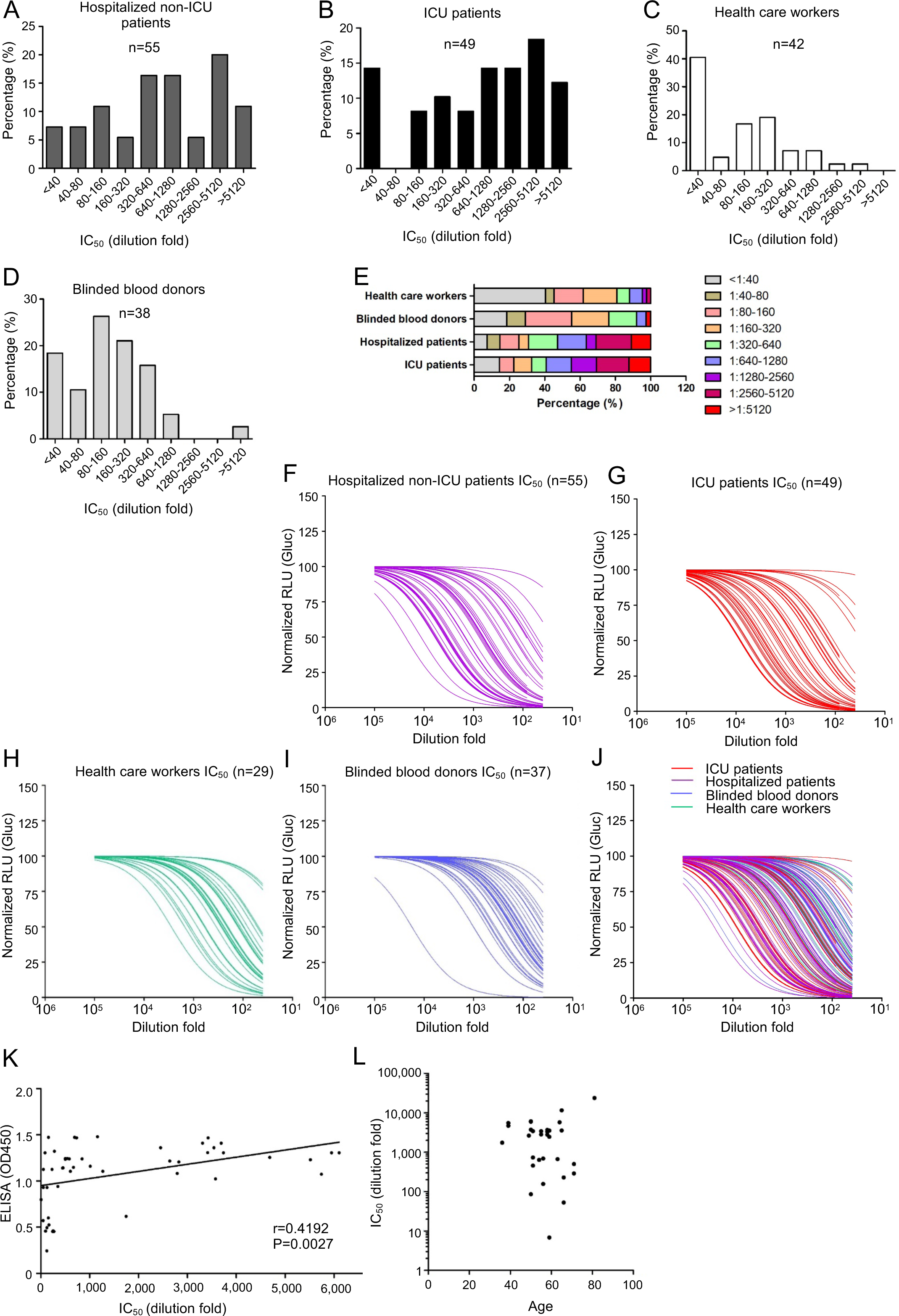
Evaluations of neutralizing antibody levels in COVID-19 hospitalized inpatients, ICU patients, health care workers, and convalescent plasma donors. Blinded serum samples were serially diluted and tested for neutralizing activity against lentiviral pseudotypes bearing SARS-CoV-2 S. **(A-E)** Ranges of neutralizing antibody titer IC_50_ in the 4 indicated groups (X axis); percent in each study group was plotted (Y axis). **(F-J)** Neutralization curves of 4 different groups, as presented by the relative infectivity of SARS-CoV-2 S pseudotypes in the presence of indicated serum samples. Y axis indicates the relative viral infectivity by setting the viral infectivity without serum to 100%; X axis indicates dilution fold of serum samples. **(K)** Correlational analysis of pseudovirus neutralization IC_50_ and N protein IgG antibody ELISA OD_450_ values. **(L)** Correlational analysis between pseudovirus neutralization IC_50_ and age.

Neutralization curves of all 4 groups of samples are presented in **Fig. 5F-J**, which again illustrates highly diverse levels of S pseudotype virus neutralizing antibody among these samples. By excluding those having an ELISA OD_450_ cut-off value of 0.40 or below (except one which is 0.24 from a PCR-positive HCW), we obtained a good correlation (r=0.4192, p < 0.0027) between the S pseudotype virus neutralization titer and the ELISA OD_450_ values (**Fig. 5K**), the latter of which was for the N protein of SARS-CoV-2 *(36)*, indicating that the N-based ELISA assay can serve as a reliable method for large-scale screening of antibody responses to SARS-CoV-2 infection. Of note, there was no correlation between levels of neutralizing antibody with age (**Fig. 5L**).

### Creation of a secreted Nano-Luciferase (secNluc)-based lentiviral vector with improved stability and sensitivity for measuring SARS-CoV-2 neutralization

While highly sensitive and convenient, one disadvantage of using Gluc as a reporter gene compared with firefly or other luciferase forms is the rapid decay of its signal during measurement. To alleviate this complication, we created a secreted Nano-Luciferase (secNluc)-based intron HIV-1-NL4.3 vector, in which the Gluc gene in the inGluc HIV-1 vector was replaced by an IL6-NanoLuc reporter, also split by an intron. Nluc is about 100-fold brighter than firefly or renilla luciferase and produces high intensity, glow-type luminescence; having an IL6 which contains a secretive signal at its N-terminus would render this reporter to be secreted into culture medium as is Gluc. We tested this new vector with SARS-CoV-2 S pseudotyping, and found that, as expected, the secNluc signal was much more stable, showing no apparent loss in signal over the 1 h testing period. In contrast, the inGluc signal decayed rapidly, within a matter of minutes (**Fig. 6A**). Importantly, this new vector was pseudotyped by SARS-CoV-2 S as efficiently as that for the inGluc vector, yet exhibited a significantly more robust and increased luciferase signals. Even at 24 h post-infection, the secNluc signal of SARS-CoV-2 S pseudotypes were about 100-fold above the background level (**Fig. 6B and C**), which shortened the detection window from the prior 48 h.

**Figure 6.**
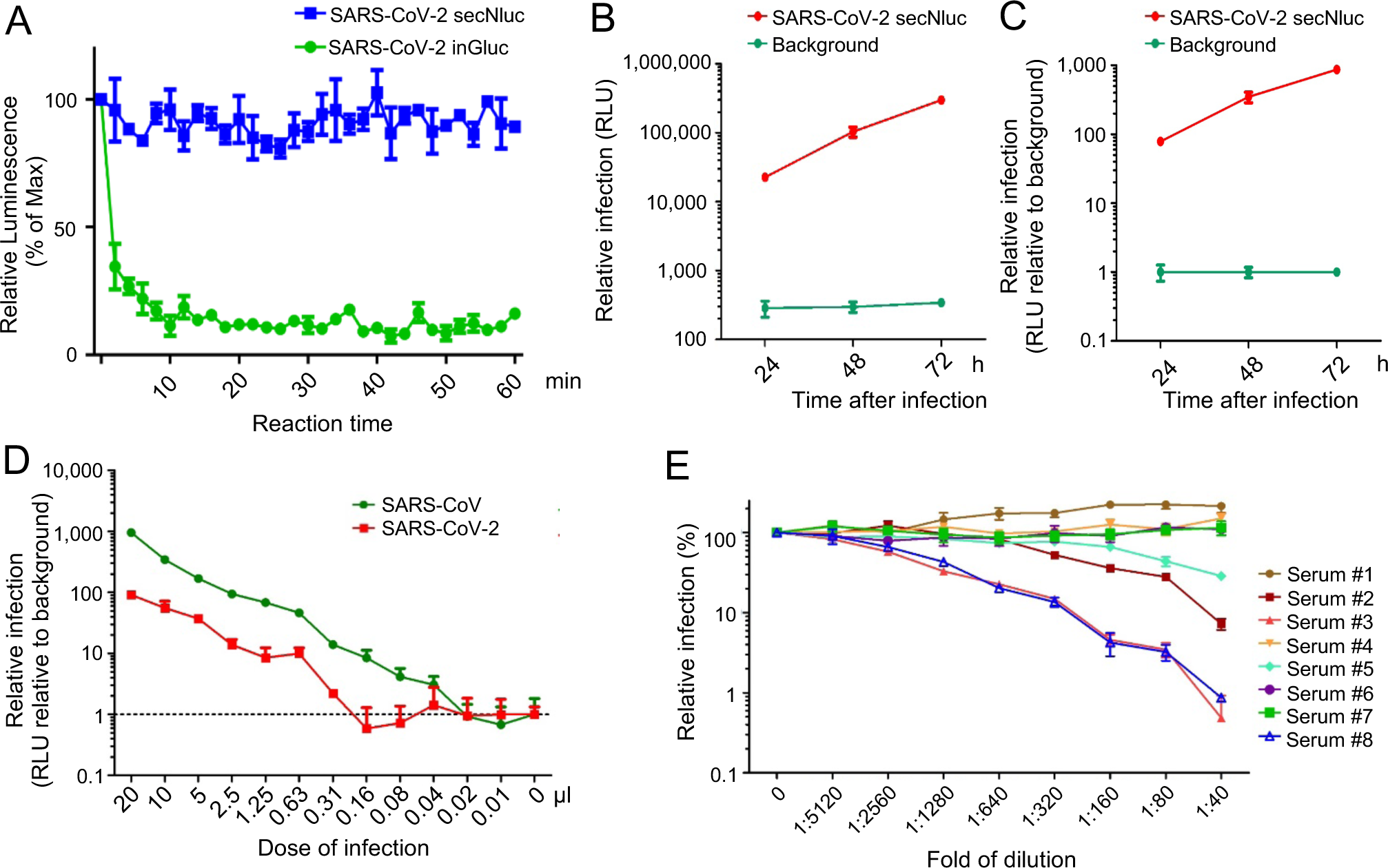
A secreted NanoLuc-based lentiviral SARS-CoV-2 S neutralization assay with improved stability and sensitivity and its application in measuring SARS-CoV-2 neutralizing antibody levels in COVID-19 patients. 293T cells were transfected with lentiviral vector (pNL4-3 inGluc or pNL4-3 secNluc) along with the SARS-CoV-2 S-C9 plasmid. Media was 48 hrs after transfection and used to infect 293T/ACE2 cells; luciferase activity was measured at indicated times to determine the viral infectivity. **(A)** Stability of inGluc and secNanoLuc luciferase signals measured over time. 20 μl of Gaussia luciferase substrate or 20 μl of Nano Luciferase substrate were added simultaneously, and luminescence measurements were then read every 2 minutes for 60 min. Plotted are the luminescence reads relative to the 0 min time point, which was set to 100 percent; secNluc exhibited a signal that was more stable than the inGluc virus infected cells. (**B and C**) Infectivity of SARS-CoV-2 S secNluc pseudotypes. **(B)** The Nano-luciferase activity of culture medium harvested from virus-infected cells at indicated times. **(C)** Relative viral infectivity was plotted by setting the mock infection to 1.0. **(D)** Indicated amounts of viral supernatant were used to infect 293T/ACE2 cells seeded in 24-well plates, and 20 μl of supernatant from virus-infected cells was used to measure the secNuc activity as shown. The dashed line indicates the background luminescence. **(E)** Experiment was performed as described in the legends of Fig. 3A and Fig. 4B, except secNluc lentiviral pseudotypes were used for infection.

To further determine assay sensitivity, we carried out infection with end-point diluted secNluc peseudotypes as we did for the inGluc vector (**Fig. 1D**), and observed that in a typical experiment as little as 0.31 ul of secNluc SARS-CoV-2 S pseudotypes showed an above-the-background level of Nluc activity at 24 h post-infection (**Fig. 6D)**, confirming that this new vector indeed shortened the SARS-CoV-2 S pseudotype virus neutralization assay to within 24 h. The estimated titers of secNluc SARS-CoV-2 and SARS-CoV S pseudotypes were ∼1.6 × 10^5^ and ∼1.3 × 10^6^ infectious units per ml, respectively, a 2-4∼fold increase compared to that from the inGluc-based system.

We next used the second generation secNluc SARS-CoV-2 S pseudotype system to measure neutralizing antibody levels of COVID-19 patient sera, which had been determined by the inGluc S pseudoypes. We observed that samples 3 and 8 still exhibited the strongest neutralizing activity, with a calculated IC_50_ of 1:4189 and 1:2556, respectively. Samples 2 and 5 also showed strong neutralizing activities, with an IC_50_ between 1:1,000 – 1:1,800. This pattern of neutralizing activity was concordant with results obtained with the inGluc assay (**Figs. 3 and 4)**. As expected, four negative control samples 1, 4, 6, and 7 showed no neutralizing activity (**Fig. 6E**). Overall, these data demonstrate that the second-generation assay, leveraging the secNluc-based SARS-CoV-2 pseudotype virus, is rapid and has a significantly improved stability and sensitivity to accurately measure the neutralizing antibody levels of COVID-19 patients.

## DISCUSSION

In this work, we described a sensitive and reliable SARS-CoV-2 S-bearing lentivirus inGluc neutralization assay that is validated by the authentic SARS-CoV-2 plaque-reduction assay. We evaluated the neutralizing antibody response in four groups of individuals with the new pseudotype virus assay: general hospitalized COVID-19 inpatients, ICU patients, university HCWs exposed to SARS-CoV-2, and convalescent plasma donors. In general, we found that hospitalized COVID-19 patients, especially ICU patients, which were PCR confirmed, had a higher neutralizing antibody titer against the SARS-CoV-2 S pseudotype virus (50% >1:640), some reaching >1:5,120, as compared with PCR-positive HCWs and convalescent plasma donors, the majority of whom had titers of <1:640. We also examined possible cross-reactivity of several reference monoclonal antibodies and polyclonal sera against SARS-CoV and SARS-CoV-2 using our new assay, and clearly ruled out cross-reactivity with the two closely related viruses. Indeed, none of the eight COVID-19 patient sera showed any neutralization of SARS-CoV S pseudotype virus. Lastly, we were able to improve the lnGluc-based SARS-CoV neutralization assay by modifying the lentiviral vector with a secNluc reporter, which makes the assay even more rapid and sensitive.

A principle of lentiviral pseudotyping, as with many other pseudotyping systems, is incorporation of viral envelope glycoproteins into the vector of interest *(37)*. In this work, by using C9-tagged S constructs, we demonstrate that the S proteins of both SARS-CoV-2 and SARS-CoV are incorporated into the HIV-1-NL4.3-inGluc vector, yet with different efficiencies. The SARS-CoV-2 S protein appears to be less efficiently incorporated compared to that of SARS-CoV, which may account in part for the relatively lower infectivity of SARS-CoV-2 S pseudotypes. This occurs despite comparable surface expression of these S proteins on viral producer cells. Another interesting observation from this study is that the C9-tagged SARS-CoV-2 S shows ∼10-fold higher titer over the WT spike, yet there is no increase in titer between C9-tagged SARS-CoV S and its WT. Work is ongoing to further decipher the underlying mechanisms of these observations. Importantly, the increased titer of SARS-CoV-2 inGluc pseudotypes bearing C9-tagged S faithfully mimic that of WT in evaluating the neutralizing antibody response in COVID-19 patients and SARS-CoV-2-exposed individuals.

The inGluc-based and secNluc-based lentiviral SARS-CoV-2 S pseudotype virus neutralizing assays have several advantages over others that have been reported *(18-22)*. First, the assays reported here use a naturally secreted Gluc, or a modified Nluc, which is secreted as a reporter; thus requiring no cell lysis or detachment of target cells before measurements of reporter gene expression *(28)*. Moreover, culture media containing the secreted Gluc or Nluc can be harvested and measured at multiple times during the infection period, which greatly increases the flexibility, efficiency and reproducibility of this assay. Second, our assay utilizes an intron in the sense genome of HIV-1 vector that splits the antisense Gluc or Nluc gene, and only after viral infection of target cells, when the intron is spliced out, can the full-length Gluc or Nluc gene be generated, leading to their expression and detection *(29, 30)*. This feature eliminates background luciferase activity possibly carried over from viral producer cells, which could otherwise confound data analyses. Third, the HIV-1-NL4.3 inGluc or secNluc vector expresses Gag-Pol in addition to accessory genes, allowing for a relatively high dose of S protein-coding plasmids to be co-transfected. This feature is particularly helpful, given the presence of a putative ER retention signal in the cytoplasmic tail of the SARS-CoV-2 S protein *(38, 39)*, which intrinsically may restrict its expression on the plasma membrane of producer cells and its incorporation into the viral particles. Lastly, this assay is simple, rapid, and cost-effective, because fewer procedures and reagents are needed compared to other reporter gene-based assays. These advantages make the inGluc- and secNluc-based virus neutralization assay particularly suitable for large-scale testing of COVID-19 serum or plasma from clinical cases, virus-exposed individuals, convalescent plasma donors and recipients, vaccinated humans and animals, as well as clinical trial participants. In addition, the new assays can be used for high-throughput screens of monoclonal antibodies and inhibitors for SARS-CoV-2, as well as many emerging viral pathogens.

## MATERIALS AND METHODS

### Constructs, reagents, and cell lines

Constructs used for production of lentiviral pseudotypes included HIV-1-NL4-3-inGluc vector *(29, 30)*, which was originally obtained from David Derse’s lab at NIH and Marc Johnson’s lab at the University of Missouri. Plasmids pcDNA3.1-SARS-CoV-S-C9, and pcDNA3.1-SARS-CoV2-S-C9 *(31)*, which encodes codon-optimized full-length spikes tagged with C9 at the C-terminus were from Fang Li’s lab at the University of Minnesota. Plasmids pcDNA-SARS-CoV-S, and paH-SARS-CoV-2-S *(40)*, which encodes codon-optimized full-length spikes were from Jason McLellan’s lab at the University of Texas-Austin. The mouse monoclonal antibody 2B04 against SARS-CoV-2 was a gift of Ali Ellebedy at Washington University *(34)*. Antibodies used for Western blotting included anti-C9 (anti-rhodopsin) (Santa Cruz, #57432), anti-p24 (abcam, #ab63917), anti-β-actin (Sigma, #A1978) and secondary antibodies anti-Mouse IgG (Sigma, #A5278), anti-Rabbit IgG (Sigma, #A9169). Secondary antibodies used for flow cytometry included FITC-conjugated anti-human IgG-Fc (Sigma,405 #F9512).

HEK293T (ATCC CRL-11268, RRID: CVCL_1926), HeLa (ATCC, CCL-2, RRID: CVCL_0030), HTX (a subclone of HT1080), A549 (ATCC, CCL-185, RRID: CVCL_0023) and Huh7.5 (RRID: CVCL_7927) cells were grown in Dulbecco’s modified Eagle’s medium (DMEM), supplemented with 1% penicillin/streptomycin and 10% (vol/vol) FBS. Calu-3 (ATCC, gift of Stelle Cormet-Boyaka) were grown in Eagle’s Minimum Essential Medium (EMEM), supplemented with 1% penicillin/streptomycin and 10% (vol/vol) FBS. HEK293T/ACE2 cell line is a gift from Dr. Fang Li at the University of Minnesota. HeLa, A549, HTX and Huh7.5 cells stably expressing ACE2 were generated by transduction of pLenti-GFP vectors expressing ACE2 (OriGene, #RC208442L4), followed by puromycin selection (1 μg/mL) for 6 days. All cell lines utilized were maintained at 37°C, 5% CO2. Authentic SARS-CoV-2 virus US-WA-1 strain was obtained from BEI Resources 418 (#NR-52281).

### Creation of a secreted intron-bearing Nano-Luc lentiviral vector

The secreted Nano-luciferase (secNluc) construct (Promega, #N1031, gift of Walther Mothes) contains the nano-luciferase construct with an N-terminal IL6 secretion signal allowing the nano-luciferase to be secreted, similarly to Gaussia Luciferase. N-terminal and C-terminal portions of this construct were cloned to include the same beta-globin intron as the Gaussia luciferase construct, also in the opposite orientation. This fragment was then introduced into the original pNL4-3 inGluc vector with the inGluc cassette removed. This produced a similar construct containing a sense orientation HIV-1 gag-pol gene and an anti-sense orientation secNluc gene containing a sense orientation beta-globin intron. This allows for the production of pseudotyped virus containing the secNluc gene without the production of secNluc in the virus producing cells, as was the case for our pNL4-3 inGluc construct.

### Patient samples and specimens

All samples were de-identified specimens from a clinical laboratory and handling of these samples were under an approved IRB protocol to Dr. Shan-Lu Liu (OSU 2020H0228). Plasma and serum were collected from hospitalized COVID-19 inpatients or ICU patients, OSU health care workers, and blinded convalescent plasma donors and analyzed in a blinded manner. Plasma was prepared using EDTA, lithium heparin, or sodium citrate anticoagulated blood samples while serum was prepared using Gold top serum-separator tubes or Red top clotting tubes. Samples were incubated for 24 hrs at room temperature to allow plasma/serum to separate. Then plasma/serum was isolated and frozen (−20°C). Samples found to be severely hemolytic were rejected.

### Production of intron-Gluc- or secNluc-based lentiviral pseudotypes bearing the S protein of SARS Coronaviruses and viral infection

For intron-Gluc- or secNluc-based pseudotyped lentiviral production, we transfected HEK293T cells with HIV-1-NL4.3 inGluc or secNluc vector plus a plasmid expressing the S protein or VSV-G in a 2:1 ratio using polyethylenimine (PEI). Supernatants were harvested at 24, 48, 72 hrs post-transfection, aliquoted and stored at -80°C. For viral infection, we added appropriate amounts of virus onto target cells and incubated plates (37°C) for 6h before changing media, then measured gaussian luciferase at 24, 48, 72 hrs after infection.

### SARS-CoV and SARC-CoV-2 S pseudotype virus neutralization assay

In the virus neutralization assay, we used 100 μl virus for each well in 96-well plates. Virus was incubated with patient or control serum or plasma, mAbs or polyclonal antisera for 1 hr at 37°C. Media was then removed from seeded HEK293T/ACE2 cells and replaced with the virus/serum or plasma mixture. The infection was allowed to proceed for 6 hrs at 37°C before changing to fresh media. *Gaussia* luciferase or Nano-Luc activity was measured at 24, 48 and 72 hrs after media change. For luciferase measurement, unless specified, 20 μl of supernatant were collected from each well and transferred to a white non-sterile 96-well plate. To each well, 20 μl of luciferase substrate was added and luminescence was immediately read by a plate reader.

### Infectious virus plaque reduction neutralization assay

Serum samples were diluted in DMEM, mixed with 80 TCID50 SARS-CoV-2, and incubated for 1 h at 37°C. Serum/virus mixtures were then used to infect confluent Vero-E6 cells for 1 h at 37°C. The serum/virus was then removed from cells and replaced with 0.3% agarose (Sigma, #A9539) in DMEM/4% FBS and incubated for 72 hrs at 37°C. Cells were then fixed with 4% paraformaldehyde in PBS and stained with 0.25% crystal violet (Sigma, #C0775) in 20% ethanol/water for visualization of plaques.

### Flow cytometry analysis of spike expression on the cell surface by soluble ACE2-hFc

HEK293T cells were transfected with 1000 ng S-C9 or WT S constructs with PEI. After 36 hrs post-transfection, cells were washed with PBS, detached with PBS containing 5 mM EDTA for 10 min, washed twice with cold PBS plus 2% FBS, and incubated with soluble ACE2-hFc proteins (10 μg/mL, a gift from Jason McLellan’s lab at the University of Texas at Austin) for 2 hrs. After three washes with PBS plus 2% FBS, cells were incubated with FITC-conjugated anti-human IgG (1:200, Sigma, #F0257) secondary antibodies for 1 hr. Cells were washed three times with cold PBS plus 2% FBS and fixed with 3.7% formaldehyde for 10 min and analyzed by flow cytometry.

### Western blotting

Western blotting was performed as previously described *(41, 42)*. In brief, cells were collected and lysed in RIPA buffer (50 mM Tris, pH 7.5, 150 mM NaCl, 1 mM EDTA, 1% Nonidet P-40, 0.1% SDS, protease inhibitor cocktail), which disrupts membrane-associated proteins. Cell lysates were clarified by centrifugation for 10 min, 12,000 x g at 4°C, and boiled at 100°C for 10 min with SDS loading buffer containing 2-Mercaptoethanol. Treated samples were resolved on 10% SDS-PAGE gels, transferred to PVDF membranes, and probed with primary antibodies.

### ELISA

ELISA was performed by using the EDI Novel Coronavirus COVID-19 N protein IgG ELISA Kit (EDI, #KT-1032) following manufacturer’s protocol. Briefly, 100 μL of 1:100 diluted serum/plasmid were added to SARS-CoV-2 antigen (nucleocapsid)-coated microplates and plates were incubated for 30 min at room temperature. Following wash steps, wells were treated with 100 μl of HRP labeled anti-human-IgG tracer antibody (EDI, #31220), incubated at room temperature for 30 min, and again washed. Then 100 μl of ELISA HRP substrate (EDI, #10020) was added, incubated for 20 min at room temperature. Finally, 100 μl stop solution (EDI, #10030) was added and absorbance at 450 nm was read with a spectrophotometric plate reader using Gen 5 software.

### Statistical Analysis

Data were analyzed as mean with standard deviation (SD). Statistical analyses were performed using GraphPad Prism 5.0 as follows: One-way Analysis of Variance (ANOVA) with Bonferroni’s post-tests was used to compute statistical significance between multiple groups for multiple comparison. A *p* values of less than 0.05 was considered significant. For calculating IC_50_, nonlinear regression of XY analyses were performed and fitted with inhibition curve. For ELISA OD_450_ and IC_50_ correlation analysis, correlation of XY analyses were performed.

## Data Availability

We agree that all data will be made available upon request.

## ACKNOWLEDGEMENTS

We thank David Derse, Marc Johnson, Fang Li, Jason McLellan, Ali Ellebedy, Walther Mothes, and Stelle Cormet-Boyaka for provision of plasmids and cell lines.We thank the NIH AIDS Reagent Program and BEI Resources for supplying important reagents that made this work possible. We thank members of The Ohio State University COVID-19 working groups for sample collections and helpful discussions. This work was supported by a fund provided by an anonymous private donor to The Ohio State University; additional support of SLL’s lab includes NIH grants R01 AI112381 and R01 AI150473. JEP was supported by Glenn Barber Fellowship. RR was supported by NIH grant R01 AI121212. JY was supported by NIH grants R01 AI130110 and The Ohio State University COVID-19 Seed Grant. RM was supported by NIH grants R01HL096376, R01HL097376,R01HL098174,R01HL081784. LS was supported by NIH NICHD grant R01 HD095881. EO wassupported by NIH R01AI134035 and R01AI130231.

## Author contributions

SLL conceived and led the project. CZ and JEP performed majority of the presented experiments. RP and SP collected patient’s samples and/or performed ELISA. PQ and YMZ produced virus and performed infections. RR, LHS and JY performed infectious virus plaque reduction assay. RR, JY, RKM, LS, EO and GL contributed to data analyses and discussion. CZ, JEP and SLL wrote the paper, which was edited by LS, GO, RKM, and JY. All authors approved the paper.

## REFERENCES

1. WHO, Coronavirus Disease (COVID-19): Situation Report, 162. World Health Organization, (2020).

2. F. Wu et al., A new coronavirus associated with human respiratory disease in China. Nature 579, 265–269 (2020).

3. A. Hachim et al., Beyond the Spike: identification of viral targets of the antibody response to SARS-CoV-2 in COVID-19 patients. MedRxiv, (2020).

4. L. L. Luchsinger et al., Serological Analysis of New York City COVID19 Convalescent Plasma Donors. MedRxiv, (2020).

5. D. F. Robbiani et al., Convergent antibody responses to SARS-CoV-2 infection in convalescent individuals. BioRxiv, (2020).

6. A. Wajnberg et al., Humoral immune response and prolonged PCR positivity in a cohort of 1343 SARS-CoV 2 patients in the New York City region. MedRxiv, (2020).

7. F. T. Cutts, M. Hanson, Seroepidemiology: an underused tool for designing and monitoring vaccination programmes in low- and middle - income countries. Tropical Medicine & International Health 21, 1086–1098 (2016).

8. S. Jiang, C. Hillyer, L. Du, Neutralizing antibodies against SARS-CoV-2 and other human coronaviruses. Trends in Immunology, (2020).

9. J. Seow et al., Longitudinal evaluation and decline of antibody responses in SARS-CoV-2 infection. MedRxiv, (2020).

10. L.-P. Wu et al., Duration of antibody responses after severe acute respiratory syndrome. Emerging Infectious Diseases 13, 1562 (2007).

11. K. Callow, H. Parry, M. Sergeant, D. Tyrrell, The time course of the immune response to experimental coronavirus infection of man. Epidemiology & Infection 105, 435–446 (1990).

12. K. Duan et al., Effectiveness of convalescent plasma therapy in severe COVID-19 patients. Proceedings of the National Academy of Sciences 117, 9490–9496 (2020).

13. Y. Cheng et al., Use of convalescent plasma therapy in SARS patients in Hong Kong. European Journal of Clinical Microbiology and Infectious Diseases 24, 44–46 (2005).

14. L. Chen, J. Xiong, L. Bao, Y. Shi, Convalescent plasma as a potential therapy for COVID-19. The Lancet Infectious Diseases 20, 398–400 (2020).

15. E. M. Bloch et al., Deployment of convalescent plasma for the prevention and treatment of COVID-19. The Journal of Clinical Investigation 130, 2757–2765 (2020).

16. X. Chen et al., Human monoclonal antibodies block the binding of SARS-CoV-2 spike protein to angiotensin converting enzyme 2 receptor. Cellular & Molecular Immunology, 1-3 (2020).

17. E. Seydoux et al., Characterization of neutralizing antibodies from a SARS-CoV-2 infected individual. BioRxiv, (2020).

18. F. Schmidt et al., Measuring SARS-CoV-2 neutralizing antibody activity using pseudotyped and chimeric viruses. Journal of Experimental Medicine 217 (11): e20201181 (2020).

19. K. H. Crawford et al., Protocol and reagents for pseudotyping lentiviral particles with SARS-CoV-2 Spike protein for neutralization assays. Viruses 12, 513 (2020).

20. J. Nie et al., Establishment and validation of a pseudovirus neutralization assay for SARS-CoV-2. Emerging Microbes & Infections 9, 680–686 (2020).

21. J. Hu et al., Development of cell-based pseudovirus entry assay to identify potential viral entry inhibitors and neutralizing antibodies against SARS-CoV-2. Genes & Diseases, (2020).

22. A. E. Muruato et al., A high-throughput neutralizing antibody assay for COVID-19 diagnosis and vaccine evaluation. BioRxiv, (2020).

23. F. Wu et al., Neutralizing antibody responses to SARS-CoV-2 in a COVID-19 recovered patient cohort and their implications. (2020).

24. H. Lv et al., Cross-reactive antibody response between SARS-CoV-2 and SARS-CoV infections. Cell Reports, 107725 (2020).

25. Y. Wang et al., Kinetics of viral load and antibody response in relation to COVID-19 severity. The Journal of Clinical Investigation (2020).

26. X. Ou et al., Characterization of spike glycoprotein of SARS-CoV-2 on virus entry and its immune cross-reactivity with SARS-CoV. Nature Communications 571 11, 1–12 (2020).

27. D. Pinto et al., Cross-neutralization of SARS-CoV-2 by a human monoclonal SARS-CoV antibody. Nature, 1-6 (2020).

28. A. R. Goerke, A. M. Loening, S. S. Gambhir, J. R. Swartz, Cell-free metabolic engineering promotes high-level production of bioactive Gaussia princeps luciferase. Metabolic Engineering 10, 187–200 (2008).

29. D. Mazurov, A. Ilinskaya, G. Heidecker, P. Lloyd, D. Derse, Quantitative comparison of HTLV-1 and HIV-1 cell-to-cell infection with new replication dependent vectors. PLoS Pathog 6, e1000788 (2010).

30. J. Yu et al., IFITM proteins restrict HIV-1 infection by antagonizing the envelope glycoprotein. Cell Reports 13, 145–156 (2015).

31. J. Shang et al., Structural basis of receptor recognition by SARS-CoV-2. Nature, 1–8 (2020).

32. R. Shi et al., A human neutralizing antibody targets the receptor binding site of SARS-CoV-2. Nature, 1-8 (2020).

33. W. Li et al., Angiotensin-converting enzyme 2 is a functional receptor for the SARS coronavirus. Nature 426, 450–454 (2003).

34. W. B. Alsoussi et al., A Potently Neutralizing Antibody Protects Mice against SARS-CoV-2 Infection. The Journal of Immunology (2020).

35. Food and Drug Administration. Recommendations for Investigation COVID-19 Convalescent Plasma, Current Contents as of 7/22/20 (2020).

36. A. Krüttgen et al., Comparison of four new commercial serologic assays for determination of SARS-CoV-2 IgG. Journal of Clinical Virology 104394 (2020).

37. J. Cronin, X.-Y. Zhang, J. Reiser, Altering the tropism of lentiviral vectors through pseudotyping. Current gene therapy 5: 387–398 (2005).

38. C. E. McBride, J. Li, C. E. Machamer, The cytoplasmic tail of the severe acute respiratory syndrome coronavirus spike protein contains a novel endoplasmic reticulum retrieval signal that binds COPI and promotes interaction with membrane protein. Journal of virology 81, 2418–2428 (2007).

39. E. Lontok, E. Corse, C. E. Machamer, Intracellular targeting signals contribute to localization of coronavirus spike proteins near the virus assembly site. Journal of Virology 78, 5913–5922 (2004).

40. D. Wrapp et al., Cryo-EM structure of the 2019-nCoV spike in the prefusion conformation. Science 367, 1260–1263 (2020).

41. M. Côté et al., Critical role of leucine-valine change in distinct low pH requirements for membrane fusion between two related retrovirus envelopes. Journal of Biological Chemistry 287, 7640–7651 (2012).

42. S.-L. Liu, M. I. Lerman, A. D. Miller, Putative phosphatidylinositol 3-kinase (PI3K) binding motifs in ovine betaretrovirus Env proteins are not essential for rodent fibroblast transformation and PI3K/Akt activation. Journal of Virology 77, 7924–7935 (2003).

